# Transcriptome-wide Mendelian randomisation exploring dynamic CD4+ T cell gene expression in colorectal cancer development

**DOI:** 10.1101/2025.04.15.25325863

**Authors:** Benedita Deslandes, Xueyan Wu, Matthew A. Lee, Lucy J. Goudswaard, Gareth W. Jones, Andrea Gsur, Annika Lindblom, Shuji Ogino, Veronika Vymetalkova, Alicja Wolk, Anna H. Wu, Jeroen R. Huyghe, Ulrike Peters, Amanda I. Phipps, Claire E. Thomas, Rish K. Pai, Robert C Grant, Daniel D. Buchanan, James Yarmolinsky, Marc J. Gunter, Jie Zheng, Emma Hazelwood, Emma E. Vincent

## Abstract

**Background:** Recent research has identified a potential protective effect of higher numbers of circulating lymphocytes on colorectal cancer (CRC) development. However, the importance of different lymphocyte subtypes and activation states in CRC development and the biological pathways driving this relationship remain poorly understood and warrant further investigation. Specifically, CD4+ T cells – a highly dynamic lymphocyte subtype – undergo remodelling upon activation to induce the expression of genes critical for their effector function. Previous studies investigating their role in CRC risk have used bulk tissue, limiting our current understanding of the role of these cells to static, non-dynamic relationships only.

**Methods:** Here, we combined two genetic epidemiological methods – Mendelian randomisation (MR) and genetic colocalisation – to evaluate evidence for causal relationships of gene expression on CRC risk across multiple CD4+ T cell subtypes and activation stage. Genetic proxies were obtained from single-cell transcriptomic data, allowing us to investigate the causal effect of expression of 1,805 genes across five CD4+ T cell activation states on CRC risk (78,473 cases; 107,143 controls). We repeated analyses stratified by CRC anatomical subsites and sex, and performed a sensitivity analysis to evaluate whether the observed effect estimates were likely to be CD4+ T cell-specific.

**Results:** We identified six genes with evidence (FDR-*P*<0.05 in MR analyses and H4>0.8 in genetic colocalisation analyses) for a causal role of CD4+ T cell expression in CRC development – *FADS2, FHL3, HLA-DRB1, HLA-DRB5, RPL28*, and *TMEM258*. We observed differences in causal estimates of gene expression on CRC risk across different CD4+ T cell subtypes and activation timepoints, as well as CRC anatomical subsites and sex. However, our sensitivity analysis revealed that the genetic proxies used to instrument gene expression in CD4+ T cells also act as eQTLs in other tissues, highlighting the challenges of using genetic proxies to instrument tissue-specific expression changes.

**Conclusions:** Our study demonstrates the importance of capturing the dynamic nature of CD4+ T cells in understanding disease risk, and prioritises genes for further investigation in cancer prevention research.

## Introduction

Colorectal cancer (CRC) is the third most commonly diagnosed cancer and the second leading cause of cancer-related deaths globally (1). While several lifestyle and environmental risk factors, such as obesity, alcohol consumption, and tobacco use, have been identified, the underlying biological pathways driving CRC development remain poorly understood (2). This gap in knowledge has limited the development of effective preventative or therapeutic interventions for CRC. A growing body of evidence suggests that circulating white blood cell (WBC) counts are linked to CRC risk, progression, severity, and mortality (3–7). WBC counts, commonly used in clinical practice as a marker of immune system and overall health, can be subdivided into basophils, eosinophils, lymphocytes, monocytes, and neutrophils (8). We have previously shown a potential protective effect of higher numbers of circulating lymphocytes on CRC risk (9). However, lymphocytes are a heterogeneous group of immune cells with distinct functional roles, and it remains unclear which specific subtypes mediate this protective effect. This highlights the need for further research into the underlying mechanisms.

Among lymphocytes, CD4+ T cells are well-known for their role in anti-tumour responses either through either direct action and/or by recruitment of other immune cells (10,11). These cells are strong candidates for contributing to the protective effect of lymphocyte count against CRC risk. Upon activation, CD4+ T cells undergo extensive gene expression remodelling to shape their effector function (12,13). This activation process occurs through distinct stages (Figure 1a), beginning in a resting state (0h), followed by a minimally active stage post-activation (“lowly active” [LA]), progressing through cell division (16h post-activation), post-division (40h post-activation), and culminating in the acquisition of effector functions (5d post-activation) (12). Each of these stages is characterized by unique functional and transcriptional profiles critical for immune surveillance and response.

**Figure 1.**
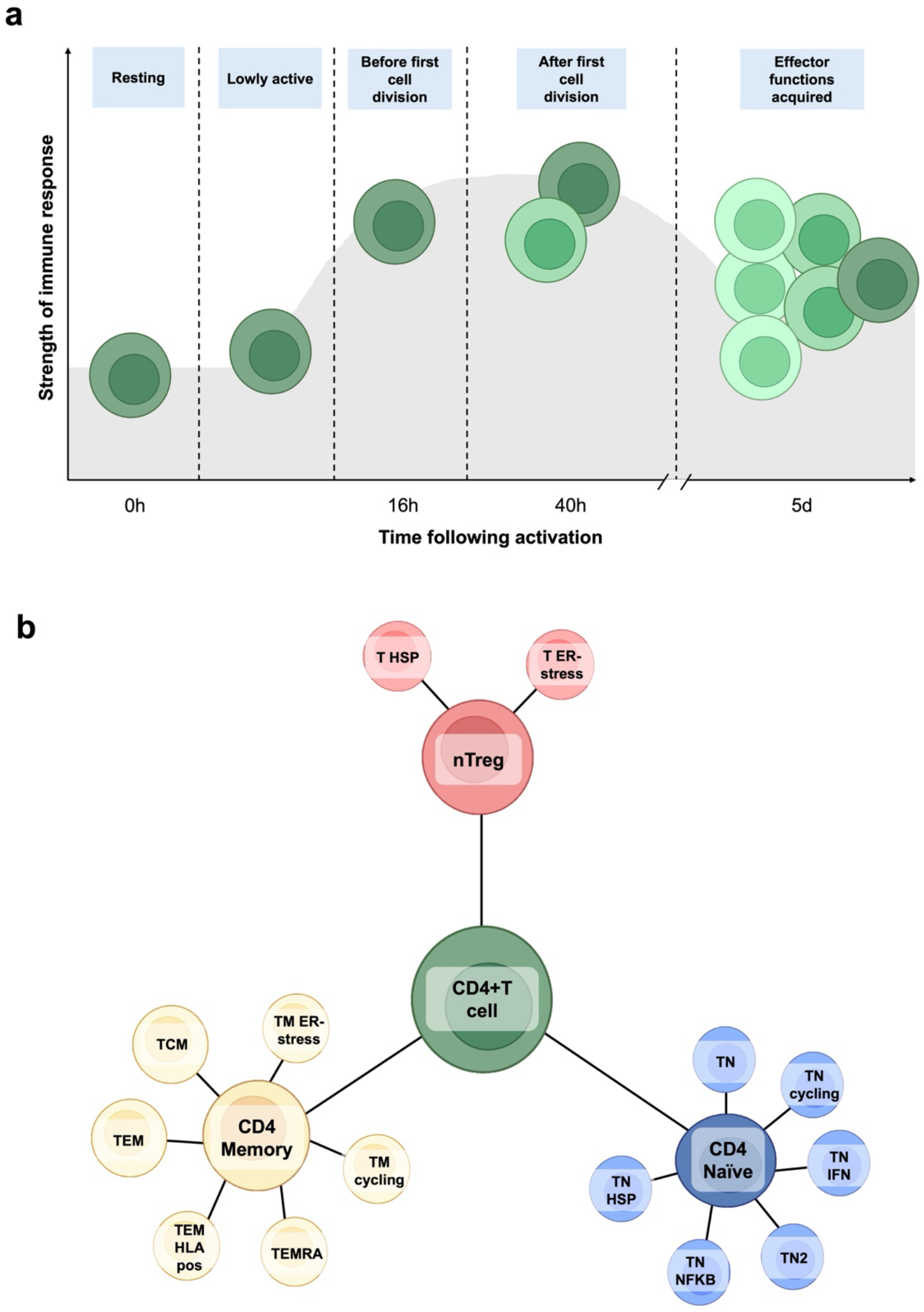
Overview of CD4+ T cell activation states and subtypes **a)** CD4+ T cells undergo complete remodelling of gene expression to shape their effector function upon activation. This activation occurs in distinct stages, progressing from a resting stage (0h), to minimally active cells following activation (“lowly active” [LA]), before undergoing cell division (16h post-activation), after completion of the first cell division (40h post-activation), and after acquiring effector functions (5d post-activation). Each activation timepoint, or state, reflects a unique functional and transcriptional profile crucial for immune surveillance and response. Colour shadings reflect transcriptomic changes. **b)** Circulating CD4+ T cell subtypes can broadly be grouped into three functional clusters: i) naïve T cells (CD4 naïve) – cells that have not yet encountered an antigen; ii) memory T cells (CD4 memory) – essential for rapid and robust responses to previously encountered antigens; iii) regulatory T cells (nTreg) – pivotal for maintaining immune homeostasis by supressing excessive immune responses.

CD4+ T cells also exhibit significant heterogeneity across their subtypes (12). Circulating CD4+ T cell subtypes can broadly be grouped into three functional clusters: i) naïve T cells (CD4 naïve) – cells that have not yet encountered an antigen; ii) memory T cells (CD4 memory) – essential for rapid and robust responses to previously encountered antigens; iii) regulatory T cells (nTreg) – pivotal for maintaining immune homeostasis by supressing excessive immune responses **(Figure 1b)** (12). Not all CD4+ T cells undergo all activation stages given some exist in a terminally differentiated state (refer to supplementary table 1 for more information). Despite their critical and distinct roles, most studies examining CD4+ T cells in CRC have relied on bulk tissue analyses, such as whole blood, which fail to capture the dynamic activation and gene expression changes of these cells. This limitation obscures the complexity of gene expression changes and hinders our ability to identify the biological mechanisms driving the protective effects of lymphocytes on CRC development.

Here, we aimed to identify key immune-related genetic drivers of CRC risk with potential to inform novel prevention or therapeutic strategies. To achieve this, we leveraged summary genetic data capturing associations between germline variants and single-cell transcriptomic data, allowing us to investigate gene expression across dynamic CD4+ T cell subtypes and activation timepoints. We performed transcriptome-wide Mendelian randomisation analyses (MR), which, under certain assumptions, can provide causal estimates (14), to identify genes that may play a role in CRC development based on single-cell transcriptomic data generated previously from CD4+ T cells (12). We then performed genetic colocalisation to evaluate possible misinference due to linkage disequilibrium and add robustness to our results. Additionally, we applied an additional sensitivity analysis to determine whether the genetic instruments used to proxy gene expression in CD4+ T cells in MR analyses are also associated with gene expression in other tissues, such as colorectal tissue. Our approach combining causal methodologies with single-cell transcriptomic data not only advances our understanding of the protective roles of CD4+ T cells in CRC risk, but also highlights potential therapeutic targets for CRC prevention.

## Methods

To identify key gene expression alterations across subtypes of CD4+ T cells with a role in CRC development, we conducted summary-level MR analyses. To evaluate the robustness of our results to misinference by linkage disequilibrium (LD), we then performed genetic colocalisation. For genes with robust evidence for an effect on CRC risk across both analyses, we performed a sensitivity analysis to evaluate the possibility of bias arising from horizontal pleiotropy through gene expression changes in other tissues. An overview of our methodology is represented in **Figure 2**.

**Figure 2.**
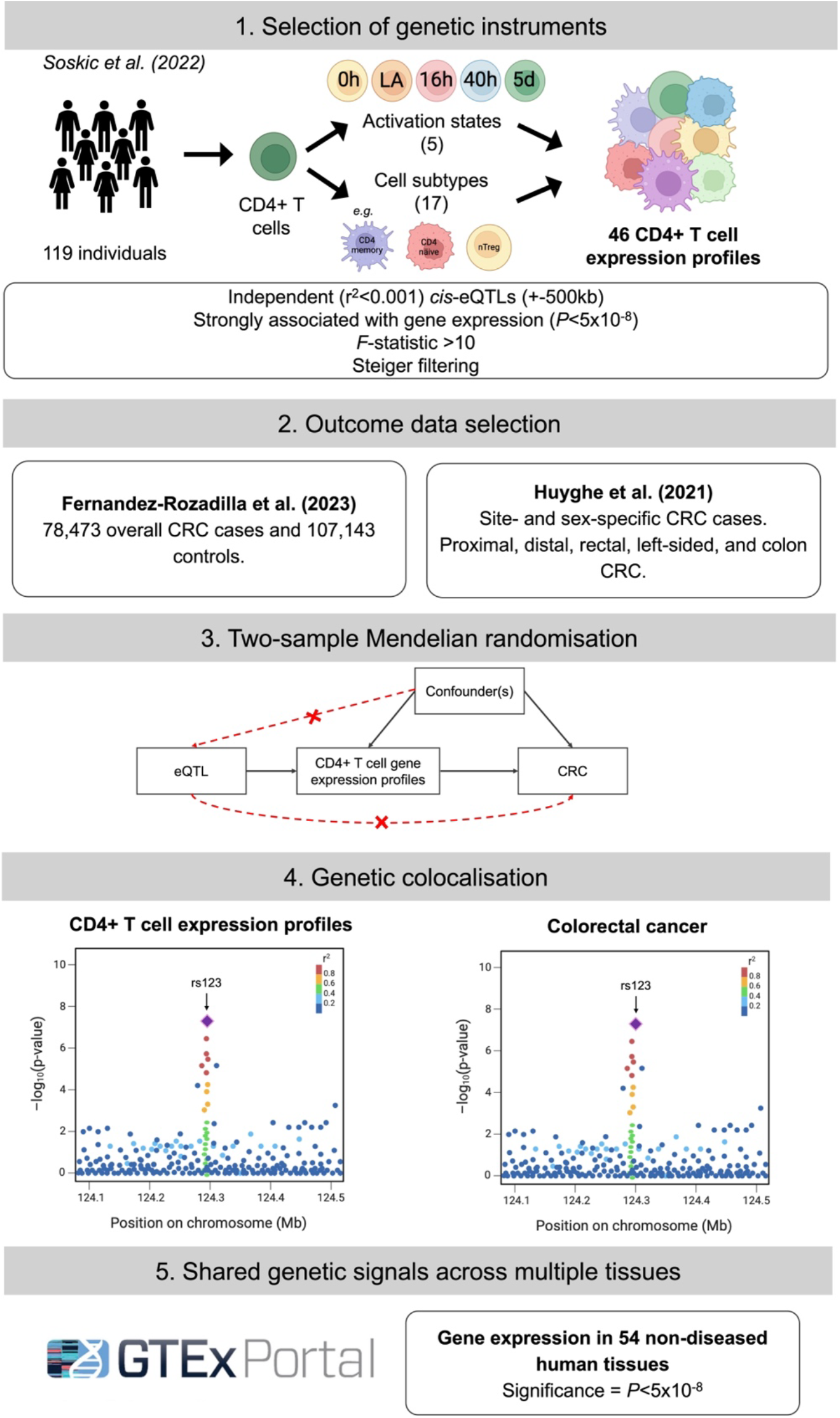
Flow chart describing study methodology. Note that genetic instruments were not available for all cell subtypes at every activation timepoint, meaning the total number of CD4+ T cell expression profiles investigated does not exactly equal the number of timepoints (5) multiplied by the number of cell subtypes (17). eQTL = expression quantitative trait loci; CRC = colorectal cancer; GTEx = Genotype-Tissue Expression.

### Study populations

#### (i) CD4+ T cell gene expression genome-wide association study (GWAS)

Soskic *et al*. isolated peripheral mononuclear cells (PMBCs) from blood samples obtained from 119 healthy individuals (56% male; 44% female) of European (“British”) ancestries (mean age 47 ± 15.61). CD4+ T cells were then further isolated from the PBMCs and activated with anti-CD3/anti-CD28 human T-Activator Dynabeads (Invitrogen) at a 1:2 beads-to-cells ratio. This resulted in a total of 655,349 CD4+ T cells across all individuals available for the single-cell transcriptomics (12).

We used summary-level expression quantitative trait loci (eQTL) results derived from single-cell transcriptomic data (12). These eQTLs were identified from the transcriptomes of CD4+ T cells sampled at five distinct timepoints following the *ex vivo* activation described above, representing a spectrum from resting to activated states **(Figure 1a)**. The timepoints included resting cells (0h), lowly active (LA), before undergoing cell division (16h post-activation), after the first cell division (40h post-activation), and after acquiring effector functions (5d post-activation) (12). Additionally, unsupervised clustering was performed by Soskic *et al*. using the transcriptomic data to group cells based on gene expression patterns and activation markers across the activation timepoints. This process identified 17 distinct CD4+ T cell subtypes which fall into three main groups – CD4 memory, CD4 naïve and nTreg **(Figure 1b)**. Combining the 17 cell types with different activation timepoints, Soskic *et al*. described a total of 46 distinct CD4+ T cell gene expression profiles (e.g. CD4_naive_0h represents CD4 naïve cells immediately following activation; note that all five activation timepoints are not available for every subtype, as they can become terminally activated before reaching the final timepoint). All CD4+ T cell subtypes included in our analyses are described in Supplementary table 1.

#### (ii) CRC risk GWAS

We used summary-level data obtained from the largest available GWAS of CRC risk in European ancestries (15) (N cases = 78,473; N controls = 107,143) and the largest available GWAS of sex-specific and site-specific CRC risk in European ancestries (16,17): proximal (14,416 cases, 43,099 controls), distal (12,879 cases, 43,099 controls), rectal (14,150 cases, 43,099 controls), left-sided (27,004 cases, 43,099 controls), colon (28,736 cases, 43,099 controls), female (24,594 cases, 23,936 controls), and male (28,271 cases, 22,351 controls). CRC classification was determined using ICD-10 codes, with the majority of cases being newly diagnosed. CRC subsites were categorized based on location: colon cancer includes the proximal colon (any primary tumour arising in the cecum, ascending colon, hepatic flexure, or transverse colon), the distal colon (any primary tumour arising in the splenic flexure, descending colon, or sigmoid colon), and cases with an unspecified site. Rectal cancer includes any primary tumour arising in the rectum or rectosigmoid junction.

### MR analyses

MR uses genetic variants, typically single nucleotide polymorphisms (SNPs), which under specific assumptions can be used in an instrumental variable framework to obtain causal estimates. The three core assumptions are: (i) the genetic variant(s) must be associated with the exposure; (ii) there are no confounders of the association between the genetic variant(s) and the outcome; and (iii) the genetic variant(s) is/are only associated with the outcome via an association with the exposure (14). In addition to these core assumptions, additional assumptions, such as exposure and outcome data being obtained from non-overlapping populations from the same underlying population in summary-level MR, also exist (18); see Sanderson *et al*. (19) for a detailed overview of MR assumptions.

We identified *cis*-eQTLs for each gene as SNPs within the gene coding region (± 500kb) which had a *P*<5×10^−8^ and were independent of other associated SNPs within a 10kb window using a linkage disequilibrium (LD) *r*^2^<0.001. We excluded weak instruments using an *F*-statistic<10 (20) and performed Steiger filtering (21) to exclude SNPs which may explain more variance in the exposure than the outcome, in order to avoid bias from potential reverse causation. As such, we identified 10,994 *cis*-SNPs associated with expression of 1,805 genes across the 46 CD4+ T cell gene expression profiles. For all MR analyses we used the Wald ratio to obtain causal estimates and the delta method to approximate standard errors (22). Benjamini-Hochberg correction (<0.05) was applied as a false discovery rate (FDR)-correction (23).

This manuscript was written following the STROBE-MR guidelines (24,25). A completed STROBE-MR checklist is included in the supplementary information.

### Genetic colocalisation analyses

Genetic colocalisation uses GWAS summary statistics to distinguish between distinct causal variants underlying a shared causal signal at a specific locus for two (or more) traits (26). Genetic colocalisation evaluates the posterior probability of five mutually exclusive scenarios: H0: there are no variants in the given genomic region causal to either trait; H1: there is a causal variant in the given genomic region for the first but not second trait; H2: there is a causal variant in the given genomic region for the second but not the first trait; H3: there is a causal variant in the given genomic region for both traits, but this variant is different between the traits; and H4: the causal variant in the given genomic region is the same for both traits (26,27).

To evaluate the possibility of misinference due to LD in our MR analyses, we performed pair-wise conditional colocalisation (PWCoCo) for all genes meeting our predetermined threshold (FDR-*P*<0.05) in MR analyses. Briefly, PWCoCo performs conditional and joint multi-SNP analysis (GCTA-COJO) to detect independent associations within a region. To achieve this, PWCoCo conditions each SNP on the sentinel SNP to identify conditionally independent SNPs for which colocalisation is then performed (28). Thus, in contrast with other genetic colocalisation methods, this approach retains the single causal variant assumption but allows for the testing of multiple causal variants within a genomic region. By combining our MR analyses with PWCoCo, we were therefore able to evaluate whether MR evidence was likely being driven by LD between distinct causal SNPs for gene expression and CRC risk; a possible violation of the third core MR assumption. Colocalisation was performed using PWCoCo for all SNPs within ±500kb of the gene coding region using prior probabilities (p1=p2=1×10^−5^ and p12=1×10^−7^) based on ∼1,621 SNPs present within each window (suitable prior probabilities chosen based on an online calculator, see reference (29) and supplementary figure 1). We interpreted posterior probabilities as a scale of evidence for a shared causal variant and set a threshold of H4>0.8 as supporting evidence for colocalisation.

### Shared genetic signals across multiple tissues

We evaluated whether evidence for a causal effect of gene expression on CRC risk from our MR and colocalisation analyses were specific to CD4+ T cells or whether the eQTLs used as genetic proxies may also instrument gene expression in other tissues. Where genetic instruments are associated with expression of the relevant gene in multiple tissues, this could suggest possible bias in our MR results from horizontal pleiotropy, a violation of the exclusion restriction assumption (i.e. the pathway from genetic instrument to CRC risk would not be through the presumed exposure of gene expression in CD4+ T cells specifically). This would be particularly pertinent if an eQTL was also associated with expression of the gene in the colon tissue itself, given the likely importance of local gene expression in disease development. To investigate this, we obtained summary statistics from the Genotype-Tissue Expression Program (GTEx) (30) for tissue-specific gene expression for all prioritised genes (i.e. those with FDR-*P*<0.05 in MR analyses and H4>0.8 in genetic colocalisation analyses). GTEx is a comprehensive public resource that maps how genetic variation influences gene expression across 54 non-diseased human tissues, based on samples from nearly 1,000 cadavers (sample size varies by tissue; see (30) for more information). We evaluated whether there was evidence for an association between the genetic instruments and expression of the instrumented gene in any of the available tissues in GTEx. We defined evidence of an association as genome-wide significance (P<5×10^−8^). Where the genetic instrument used in MR analyses was not available in the GTEx dataset, we instead evaluated the SNP in highest LD with the eQTL which was available in GTEx data (minimum required r^2^ = 0.8).

## Results

### Mendelian randomisation analyses

Results are given as odds ratio (OR) of CRC risk per standard deviation (SD) higher expression of the gene in CD4+ T cells. Of 1,805 genes (across the 46 CD4+ T cell gene expression profiles), expression of 61 genes had evidence (FDR-*P*<0.05) for a potential causal effect on CRC risk **(Figure 3**; Supplementary table 2). Among these, the activation state and cellular subtype that contributed the most were CD4+ T cells 40h post-activation **(Figure 4a)** and CD4 naïve respectively **(Figure 4b)**.

**Figure 3.**
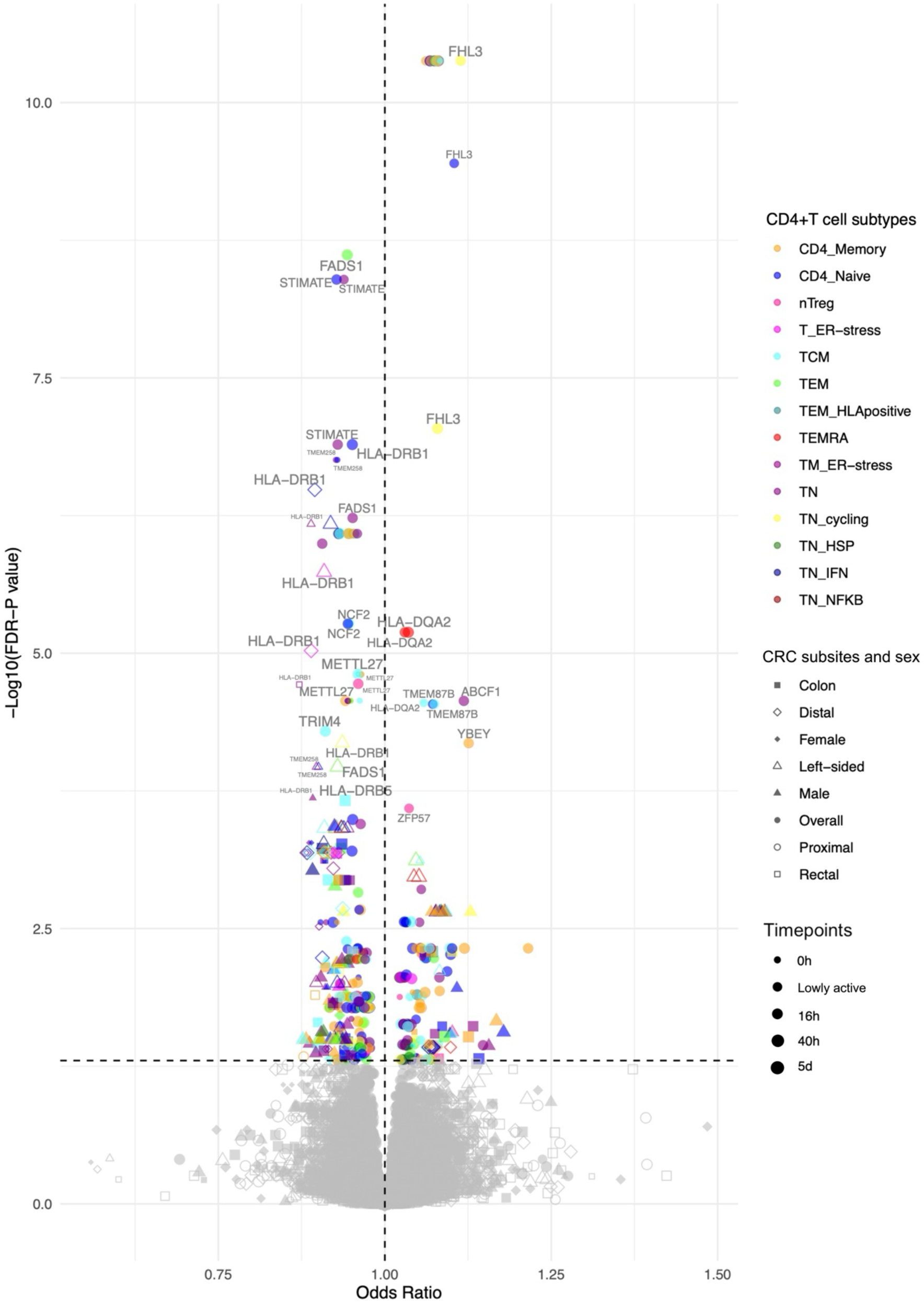
Volcano plot showing two-sample Mendelian randomisation results. Colours represent the different CD4+ T cell subtypes, shapes represent CRC subsites and sex, and point size represents activation state. Points labelled with gene names. Refer to supplementary table 1 for CD4+ T cell subtype definitions and abbreviations.

**Figure 4.**
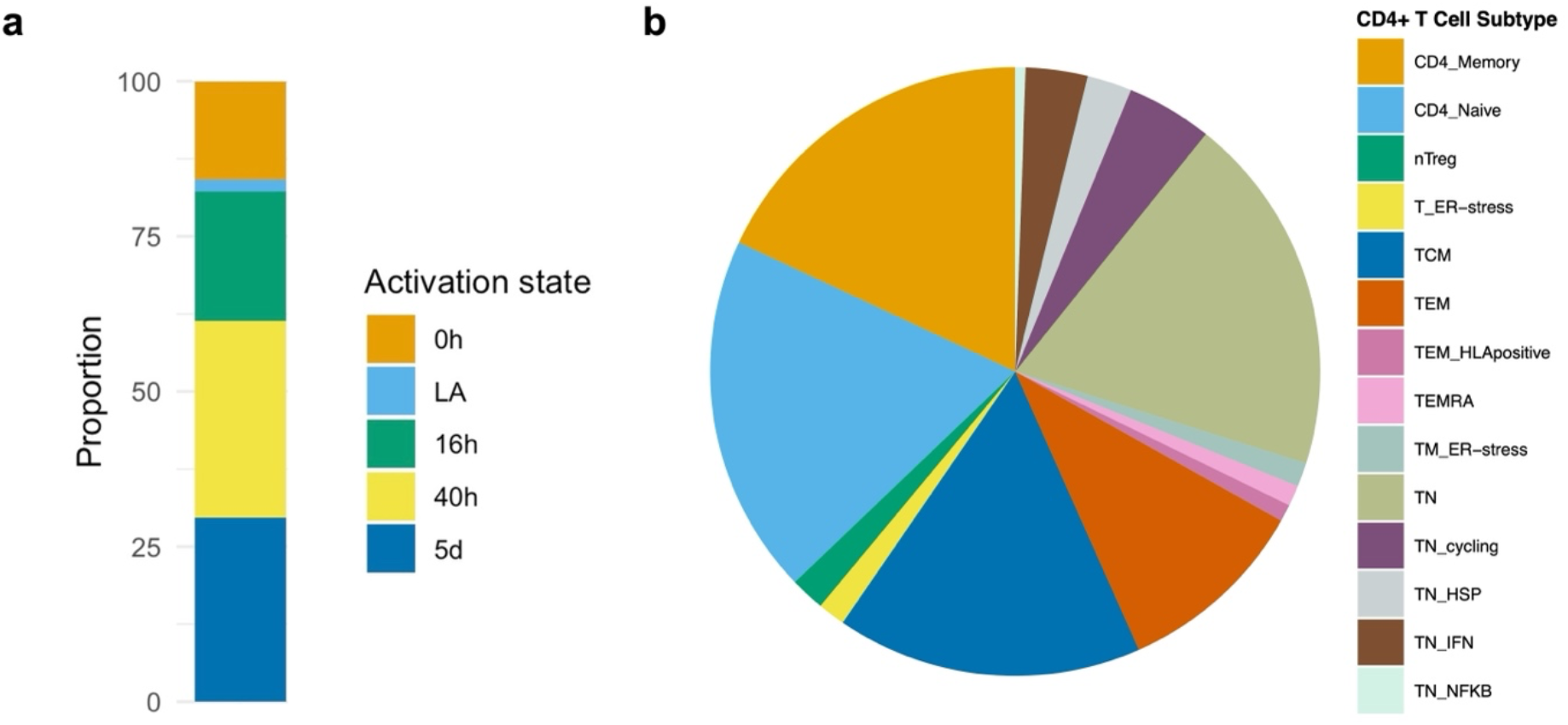
Proportion of CD4+T cell activation states and cell subtypes contributing to significant Mendelian randomisation results. **a)** Stacked bar plot representing proportion of CD4+ T cell activation states (0h, LA, 16h, 40h, 5d) contributing to main study. **b)** Pie chart representing proportion of CD4+ T cell subtypes contributing to main study results. Refer to supplementary table 1 for CD4+ T cell subtype definitions and abbreviations. LA = lowly active.

### Genetic colocalisation analyses

Of the 61 genes with evidence for an association between their expression and CRC risk, six had evidence for a shared signal with CRC risk (H4>0.8): *FADS2, FHL3, HLA-DRB1, HLA-DRB5, RPL28, and TMEM258* (Table 1; full results in supplementary table 4).

**Table 1.**
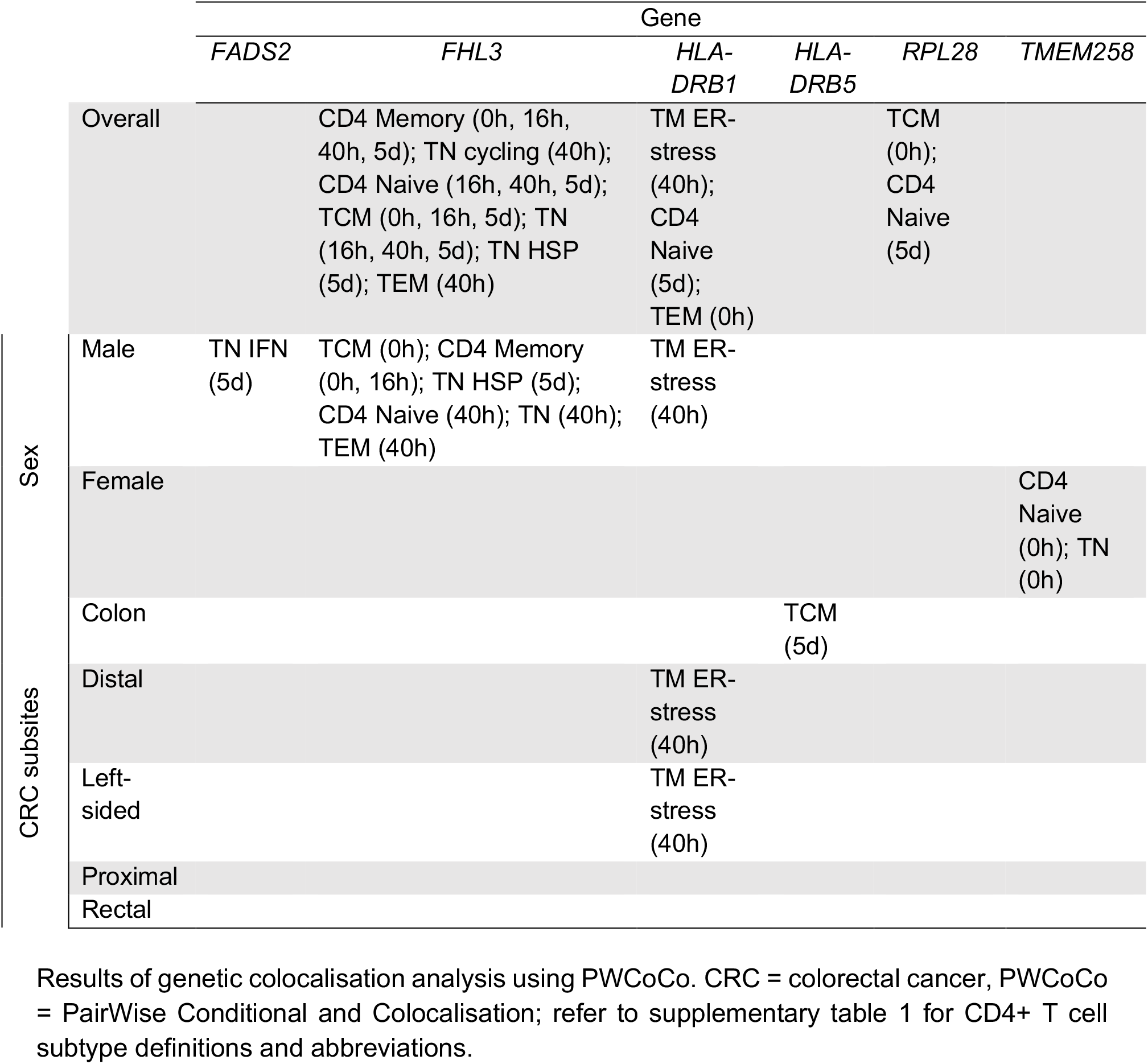
Genetic colocalisation results.

### Shared genetic signals across multiple tissues

For the six genes with robust evidence from MR (FDR-*P*<0.05) and genetic colocalisation (H4>0.8) analyses, we evaluated whether these observations could be explained through gene expression in tissues other than CD4+ T cells. Table 2 summarises the results from this sensitivity analysis.

**Table 2.**
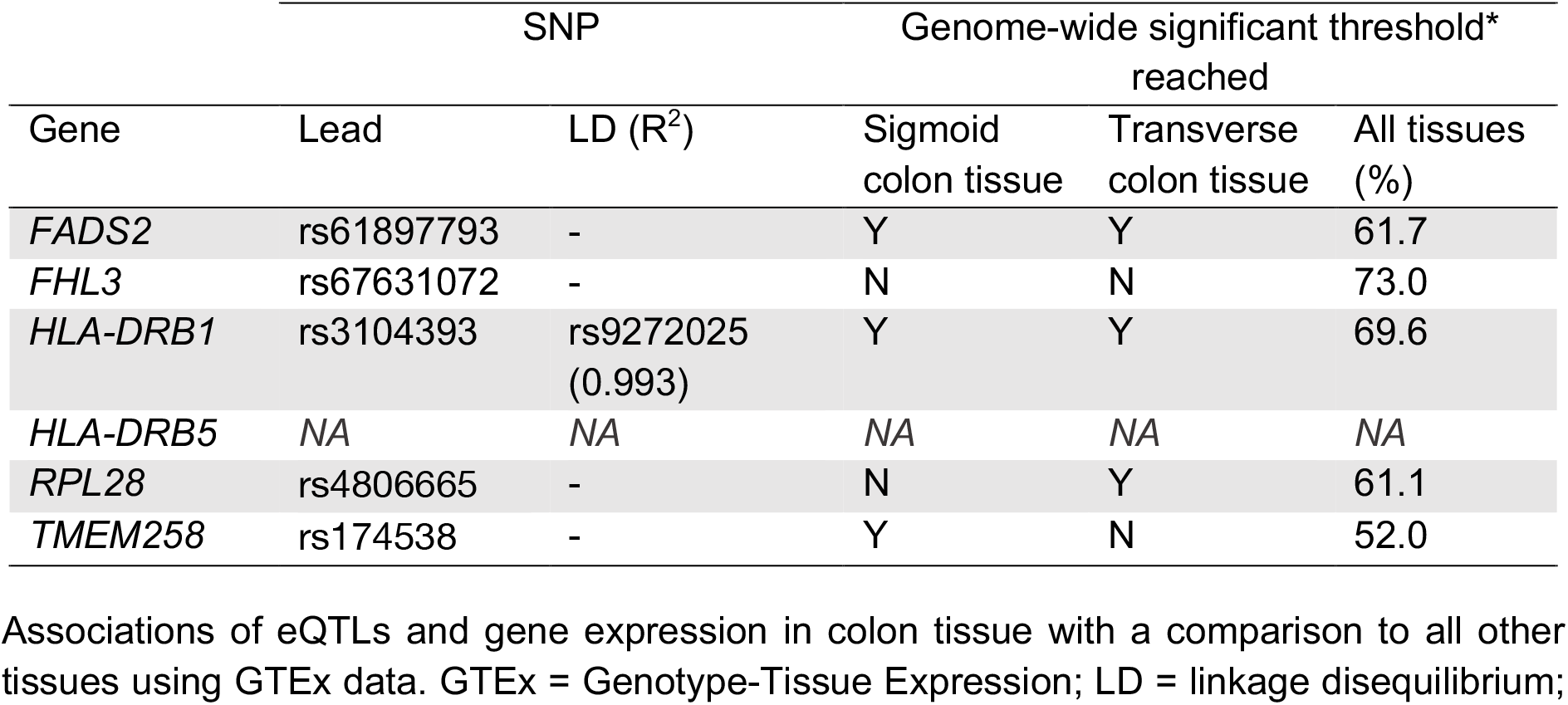
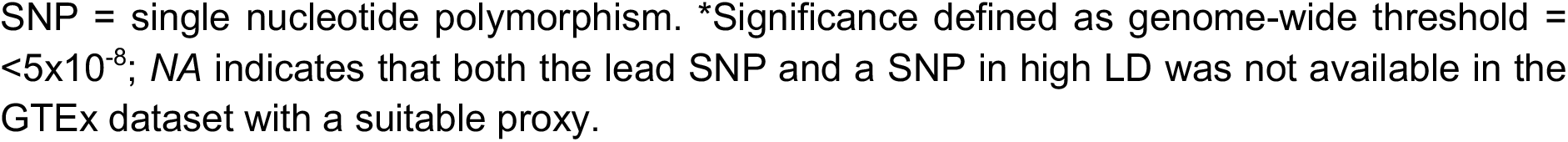
Shared genetic signals across multiple tissues.

|  |  | SNP | Genome-wide significant threshold* reached |  |  |
| --- | --- | --- | --- | --- | --- |
| Gene | Lead | LD (R<sup>2</sup>) | Sigmoid colon tissue | Transverse colon tissue | All tissues (%) |
| *FADS2* | rs61897793 | - | Y | Y | 61.7 |
| *FHL3* | rs67631072 | - | N | N | 73.0 |
| *HLA-DRB1* | rs3104393 | rs9272025 (0.993) | Y | Y | 69.6 |
| *HLA-DRB5* | NA | NA | NA | NA | NA |
| *RPL28* | rs4806665 | - | N | Y | 61.1 |
| *TMEM258* | rs174538 | - | Y | N | 52.0 |

Associations of eQTLs and gene expression in colon tissue with a comparison to all other tissues using GTEx data. GTEx = Genotype-Tissue Expression; LD = linkage disequilibrium;
SNP = single nucleotide polymorphism. \*Significance defined as genome-wide threshold = $<5 \times 10^{-8}$ ; NA indicates that both the lead SNP and a SNP in high LD was not available in the GTEx dataset with a suitable proxy.

The lead SNP and any SNPs in high LD for *HLA-DRB5* expression were not available in GTEx, meaning we were unable to include this gene in this sensitivity analysis. Overall, the lead SNPs for all five other genes with robust evidence for a role in CRC risk were associated with gene expression in other tissues in the GTEx dataset (*P*<5×10^−8^). The proportion of tissues in which eQTLs were associated with gene expression ranged from 52-73% (mean=64%), making it difficult to decipher the relevant tissue through which the identified genes act to influence CRC risk. For three of these genes (*FADS2, HLA-DRB1, RPL28*), the tissues where the eQTL associated with gene expression included transverse and sigmoid colon, highlighting a biologically plausible mechanism of horizontal pleiotropy in our analyses.

## Discussion

We used a causal framework employing MR and genetic colocalisation to investigate whether CD4+ T cell subtype- and activation timepoint-specific gene expression may have a causal role in CRC risk. We identified six genes (*FADS2, FHL3, HLA-DRB1, HLA-DRB5, RPL28*, and *TMEM258*) with robust evidence for a causal role of expression in CD4+ T cells in CRC risk. Notably, *TMEM258* has not been previously linked to CRC. Furthermore, several of these observations were specific to different CD4+ T cell activation states (e.g., prior to antigen presentation). However, the lead genetic variants associated with these six genes in CD4+ T cell subtypes at different activation points were also associated with expression in multiple other tissues, for some genes including colon tissues, suggesting the effect of gene expression on CRC risk might not be entirely specific to CD4+ T cells.

We observed evidence for sex-specific effects of gene expression in our analyses **(Figure 5 and supplementary table 3)**. For example, MR results suggested a protective effect of higher *TMEM258* expression on female-specific CRC risk in CD4 naïve cells (OR= 0.89, confidence interval (CI)= 0.85 to 0.93) and TN cells at rest (OR= 0.89, CI= 0.85 to 0.93) but not on male-specific CRC risk. *TMEM258* is involved in protein synthesis, folding and trafficking (31). Previous research has demonstrated that dysregulation of *TMEM258* expression can lead to endoplasmic reticulum (ER) stress, consequently triggering activation of the unfolded protein response (UPR) (31). UPR is known to be beneficial to CD4+ T cells as it supports differentiation, activation, cytokine production and autophagy (32). This may explain the potential mechanism by which increased *TMEM258* expression could reduce CRC risk. Additionally, we observed sex-specific effects for *FADS2*, as its expression was significantly protective against male-specific CRC development. *FADS2*, which encodes the enzyme fatty acid desaturase 2, plays a crucial role in the biosynthesis of polyunsaturated fatty acids (PUFAs), including omega-3 and omega-6 fatty acids (33). While omega-3 PUFAs have been shown to have protective associations against various cancers, including CRC, the relationship between omega-6 PUFAs and CRC risk remains unclear (34–38). We found evidence for a protective effect of *FADS2* expression on male CRC risk in naïve T cells producing interferon-gamma (IFN) upon activation (TN IFN) five days post-activation (OR=0.89, CI=0.85 to 0.93). Metabolic reconstruction, mediated by gene expression changes, is an important aspect of CD4+ T cell activity (39,40). Moreover, we observed null results for an effect estimate of *FADS2* expression in the same cell type 40 hours post-activation **(Figure 5)**, which supports an activation timepoint-specific effect of this gene and demonstrates the importance of considering the dynamic nature of CD4+ T cells.

**Figure 5.**
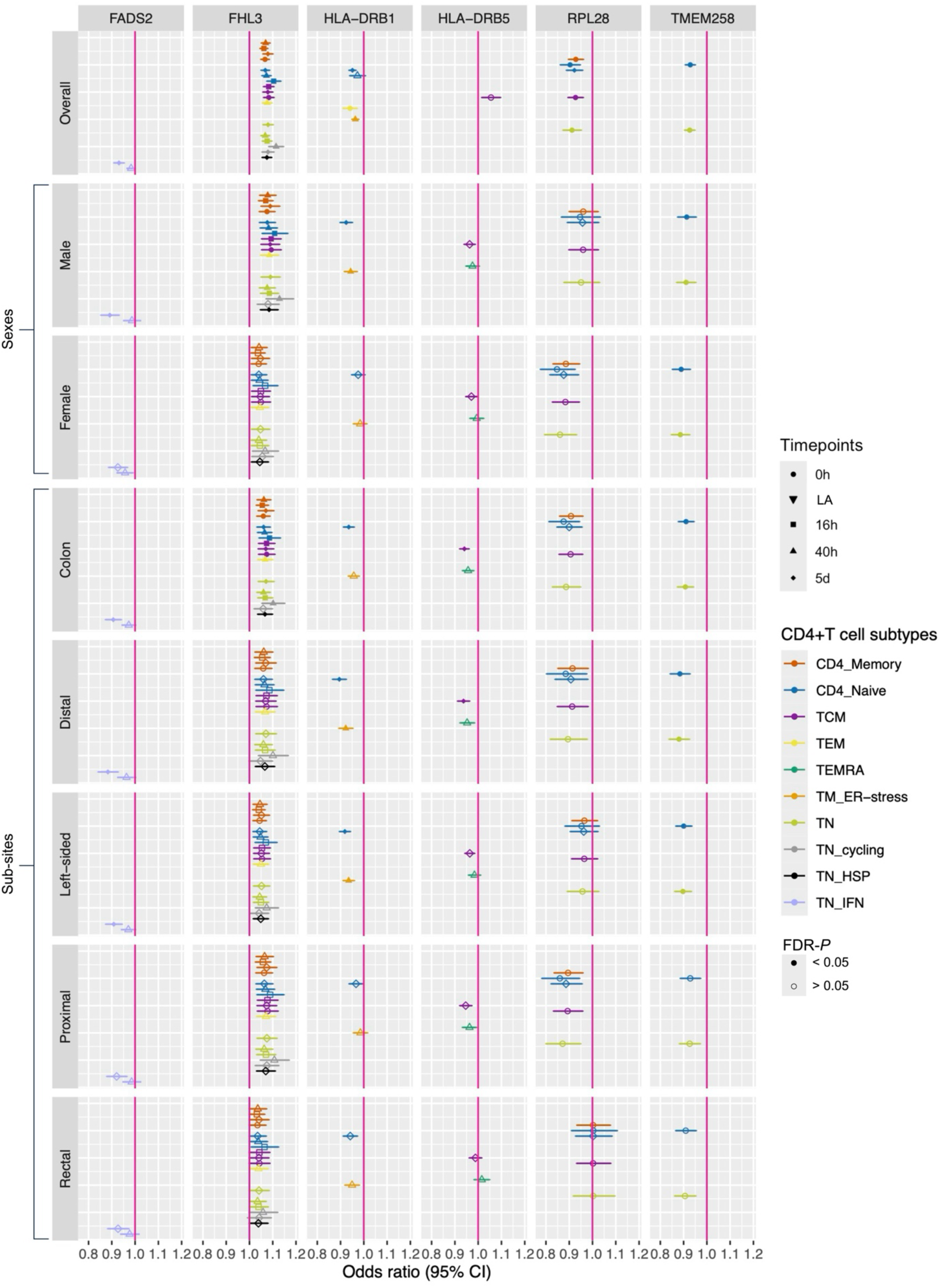
Mendelian randomisation results for genes that showed strong evidence of genetic colocalisation, separated by gene, sex and CRC subsites. Colours represent CD4+ T cell subtypes; shapes represent activation timepoint; and point fill represents whether FDR-*P* is < or > than 0.05. CRC = colorectal cancer; LA = lowly active.

In addition, we found evidence for a protective effect of two genes involved in the human leukocyte antigen (HLA) complex, *HLA-DRB1 and HLA-DRB5*, in CRC risk. We found evidence for a causal effect of higher *HLA-DRB1* expression on overall, male, distal and left-sided CRC risk in several subtypes of CD4+ T cells, particularly memory T cells experiencing ER stress (TM ER-stress) at 40h post-activation **(Figure 5)**. Cancer of the distal colon, which is part of left-sided CRC, is disproportionately diagnosed in males, which reflects these results. HLA-DRB1 is a class II major histocompatibility complex (MHC) protein involved in aiding the immune system distinguishing the body’s own proteins from those made by foreign bodies (41). The HLA-DRB1 chain is a component of the MHC class II complex on the surface of antigen-presenting cells and is responsible for antigen presentation to CD4 T helper cells, thus supporting immune activation in response to peptides from pathogens (41). Our results suggest that the beneficial effect we observe of increased expression of this gene on CRC may be related to enhanced antigen presentation to CD4+ T cells, consequently boosting anti-tumour immunity. We found strong evidence for a protective effect of *HLA-DRB5* expression on colon-specific CRC risk in memory T cells (TCM) five days post-activation (OR= 0.94, CI= 0.92 to 0.96). Similar to *HLA-DRB1, HLA-DRB5* is an MHC class II protein (41). However, unlike *HLA-DRB1, HLA-DRB5* is only expressed in a subset of the population (42). This protein is involved in presenting peptides to T-helper cells, aiding in the initiation of an immune response (41). These findings reinforce the importance of immune vigilance/surveillance and its protective effect against CRC development (43). Higher *HLA-DRB5* expression may also enhance antigen presentation, resulting in stronger and more specific anti-tumour responses. Our results **(Figure 5)** show that higher *HLA-DRB5* expression in TCM cells 5 days post-activation has a protective effect risk of colon cancer, which aligns with this explanation. While we observed evidence for a protective effect of *HLA-DRB5* expression on colon-specific CRC risk in TCM cells, key players in long-term immune memory and response (44), we found little evidence for similar effect in effector memory cells re-expressing CD45RA (TEMRA). This highlights a cell type-specific relationship and underscores the importance of considering distinct subtypes of CD4+ T cells.

We observed a protective effect of higher *RPL28* expression on overall CRC risk in TCM cells at rest (OR = 0.92, CI = 0.89 to 0.96) and CD4 naïve cells 5 days post activation (OR = 0.89, CI = 0.82 to 0.96). The *RPL28* gene encodes for a ribosomal protein component of the large ribosome subunit (60S) and is involved in the assembly of ribosomes and the translation of messenger ribonucleic acid (mRNA) into proteins (45). As TCM cells play an important role in long-term immune memory and surveillance, *RPL28* may facilitate the production of necessary proteins that contribute to their maintenance and longevity. Similarly, CD4+ naïve T cells at 5 days post-activation require increased protein synthesis in order to support rapid proliferation and differentiation (12).

Finally, we identified strong evidence for a detrimental effect of higher *FHL3* expression on both overall and male-specific CRC risk across several subtypes of CD4+ T cells during rest and at three post-activation timepoints **(Figure 5)**. *FHL3* encodes the Four and a Half LIM Domains 3 (FHL3) protein, which is highly expressed in skeletal muscle. Although its specific function in this tissue is unclear (46), it is believed that the FHL family being localized to the nucleus, plays a critical role in transcription regulation (47). Differential expression of FHL3 could influence downstream gene expression. *FHL3* has been previously identified as a potential susceptibility gene for CRC, with abnormal expression patterns observed in several cancers. However, the direction of its association varies by cancer type (47–52). Given these findings, further research is necessary to elucidate the mechanisms underlying the detrimental effects suggested by our results.

Our study combined two complimentary methods which, taken together, provide evidence for a causal relationship that is robust to biases and confounding factors commonly associated with traditional epidemiological studies (14). Given the complexity and dynamic nature of CD4+ T cells, we aimed to identify genes with a role in CRC risk using single-cell data spanning a range of cell subtypes and activation points. However, several limitations to our analysis exist. First, we used data from European ancestries which, though we assume are homogenous and therefore satisfy our genetic instrument assumptions, means these results may not be generalizable to other populations. Second, sensitivity analyses revealed that genetic variants associated with gene expression in CD4+ T cells were also linked to gene expression in other tissues, suggesting that our findings may reflect broader tissue-level expression changes rather than being CD4+ T cell-specific. Third, two of the identified genes, *HLA-DRB1* and *HLA-DRB5*, are located within the MHC region on chromosome 6, a region known for high genetic variability and LD due to its complex genetic architecture. While biologically relevant, this variability may introduce biases to our study (53,54), consequently warranting cautious interpretation on findings related to these genes. Future methodologies may better resolve gene colocalisation within the MHC region (55). Fourth, though we investigated our outcome stratified by sex, it was not possible to do this for our exposures and it is unclear whether sex is an important factor in the genetic architecture of gene expression. Lastly, we acknowledge that, according to STROBE-MR guidelines, an ideal approach would include a replication dataset to validate our results, though this was not feasible given the novelty of the underlying data.

## Conclusion

Our analysis identified six genes with robust evidence for a causal effect of expression in CD4+ T cell subtypes on CRC risk, including TMEM258, a gene not previously reported in relation to CRC development. This highlights its potential as a novel candidate for further research into CRC pathogenesis. Additionally, our findings revealed significant variability in causal estimates of CRC risk across different CD4+ T cell subtypes, activation time points, CRC anatomical subsites, and sex. These observations underscore the complex, context-dependent relationships between immune system dynamics and CRC risk.

## Data Availability

All data can be found in the manuscript, in the supplementary information, or in the links provided in the references. The GWAS of overall CRC in European ancestries can be accessed using the GWAS catalogue (https://www.ebi.ac.uk/gwas/) accession no. GCST90129505. The data where the genetic instruments were extracted for all MR analyses are available on this link (https://trynkalab.sanger.ac.uk). All code used to carry out analyses has been made publicly available on GitHub (https://github.com/bennydeslandes/CD4-T_cell_CRC). Further information on the TwoSampleMR package and PWCoCo can be found on https://github.com/MRCIEU/TwoSampleMR/, and https://github.com/jwr-git/pwcoco, respectively.

## List of abbreviations

CI: Confidence interval
CRC: Colorectal cancer
eQTL: Expression quantitative trait locus
ER: Endoplasmic reticulum
FADS: Fatty acid desaturase
FDR: False discovery rate
FHL3: Four and a Half LIM Domains 3
GCTA-COJO: Conditional and joint multi-SNP analysis
GECCO: Genetics and Epidemiology of Colorectal Cancer Consortium
GTEx: Genotype-Tissue Expression
GWAS: Genome-wide association study
HLA: Human leukocyte antigens
IFN: Interferon
LA: Lowly active
LD: Linkage disequilibrium
MHC: Major histocompatibility complex
MR: Mendelian randomisation
mRNA: Messenger ribonucleic acid
nTreg: Natural regulatory T cell
OR: Odds ratio
PUFAs: Polyunsaturated fatty acids
PWCoCo: PairWise Conditional and Colocalisation
RPL28: Ribosomal protein L28
rsID: Reference SNP cluster IDs
SD: Standard deviation
SNP: Single-nucleotide polymorphism
TMEM258: Transmembrane protein 258
UPR: Unfolded protein response
WBC: White blood cell

## Declarations

### Ethics approval and consent to participate

All GWAS obtained approval from the appropriate ethical committee(s) (12,15–17).

### Availability of data and materials

#### Statistical analyses

The bulk of the analyses were performed using RStudio (version 2024.4.2.764) (56). MR analyses were performed using TwoSampleMR (version 0.5.7) (57). Data was manipulated using the following packages: arrow (version 16.1.0), biomaRt (version 2.58.0), data.table (version 1.14.10), dplyr (version 1.1.4), GenomicRanges (version 1.54.1), gwascat (version 2.34.0), gwasvcf (version 0.1.2), rtracklayer (version 1.62.0), stringr (version 1.5.1), tidyr (version 1.3.0) (58–68). Plots were created using ggrepel (version 0.9.5) and ggplot2 (version 3.4.4) (69,70). Allele frequencies were calculated using PLINK2.0 (71). Genetic colocalisation analyses were performed using Cmake (version 3.20.0) and PWCoCo (version 1.0) (28). Priors used in the colocalisation analyses were computed using link in reference (29).

## Disclaimer

Where authors are identified as personnel of the International Agency for Research on Cancer/World Health Organization, the authors alone are responsible for the views expressed in this article and they do not necessarily represent the decisions, policy or views of the International Agency for Research on Cancer/World Health Organization.

## Authors’ contributions

BD: Data analysis – conducting MR and genetic colocalisation analysis, Writing – original draft, Writing – review and edit. XW: Data analysis – selecting genetic instruments, Writing – review and edit. MAL: Visualization – creating figures, Methodology – genetic colocalisation analyses, Writing – review and edit. LJG: Writing – review and edit. GWJ: Writing – review and edit. AG – data collection. AL – data collection. SO – data collection. VV – data collection. AW – data collection. AHW – data collection. JRH – data collection. UP – data collection. AIP – data collection. CET – data collection. JY: Methodology – genetic colocalisation analyses, Writing – review and edit. MJG: Writing – review and edit. JZ: Writing – review & editing, Writing – original draft, Methodology - conducting MR and genetic colocalisation analysis, Supervision, Project administration, Methodology and conceptualization. EH: Writing – review & editing, Writing – original draft, Methodology - conducting MR and genetic colocalisation analysis, Supervision, Project administration, Methodology and conceptualization. EEV: Writing – review & editing, Writing – original draft, Supervision, Project administration, Methodology and conceptualization.

## Acknowledgements

This work used the computational facilities of the Advanced Computing Research Centre, University of Bristol - http://www.bristol.ac.uk/acrc/. The authors would like to acknowledge the University of Bristol’s High Performance Computing (HPC) team, without whom this work would not have been possible.

