## Supplementary note for "Transcriptome-wide Mendelian randomisation exploring dynamic CD4+ T cell gene expression in colorectal cancer development"

**STROBE-MR checklist of recommended items to address in reports of Mendelian randomization studies**^1^ ^2^

| **Item No.** | **Section** | **Checklist item** | **Page No.** | **Relevant text from manuscript** |
| --- | --- | --- | --- | --- |
| 1 | **TITLE and ABSTRACT** | Indicate Mendelian randomization (MR) as the study’s design in the title and/or the abstract if that is a main purpose of the study | 1 | Title: “Transcriptome-wide Mendelian randomisation exploring dynamic CD4+ T cell gene expression in colorectal cancer development”  Abstract: “We performed two genetic epidemiological methods, Mendelian randomisation (MR) and genetic colocalisation (…)” |
|  | **INTRODUCTION** |  |  |  |
| 2 | **Background** | Explain the scientific background and rationale for the reported study. What is the exposure? Is a potential causal relationship between exposure and outcome plausible? Justify why MR is a helpful method to address the study question | 4 | Yes, justified in the introduction. |
| 3 | **Objectives** | State specific objectives clearly, including pre-specified causal hypotheses (if any). State that MR is a method that, under specific assumptions, intends to estimate causal effects | 4 | “Here, we aimed to identify key immune-related genetic drivers of CRC risk with potential to inform novel prevention or therapeutic strategies. To achieve this, we leveraged summary genetic data capturing associations between germline variants and single-cell transcriptomic data, allowing us to investigate gene expression across dynamic CD4+ T cell subtypes and activation timepoints” |
|  | **METHODS** |  |  |  |
| 4 | **Study design and data sources** | Present key elements of the study design early in the article. Consider including a table listing sources of data for all phases of the study. For each data source contributing to the analysis, describe the following: |  |  |
|  | a) | Setting: Describe the study design and the underlying population, if possible. Describe the setting, locations, and relevant dates, including periods of recruitment, exposure, follow-up, and data collection, when available. | 5-6 | Yes, study design and underlying population described in the methodology section. |
|  | b) | Participants: Give the eligibility criteria, and the sources and methods of selection of participants. Report the sample size, and whether any power or sample size calculations were carried out prior to the main analysis | 5-6 | Yes, sample size and participant information is described in the methodology, within the study populations subsection. |
|  | c) | Describe measurement, quality control and selection of genetic variants | 6 | “We identified cis-eQTLs for each gene as SNPs within the gene coding region (± 500kb) which had a P<5x10-8 and were independent of other associated SNPs within a 10kb window using a linkage disequilibrium (LD) r2<0.001. We excluded weak instruments using an F-statistic<10 and performed Steiger filtering to exclude SNPs which may explain more variance in the exposure than the outcome, in order to avoid bias from potential reverse causation. As such, we identified 10,994 cis-SNPs associated with expression of 1,805 genes across the 46 CD4+ T cell gene expression profiles. For all MR analyses we used the Wald ratio to obtain causal estimates and the delta method to approximate standard errors. Benjamini-Hochberg correction (<0.05) was applied as a false discovery rate (FDR)-correction.” |
|  | d) | For each exposure, outcome, and other relevant variables, describe methods of assessment and diagnostic criteria for diseases | 6 | “CRC classification was determined using ICD-10 codes, with the majority of cases being newly diagnosed. CRC subsites were categorized based on location: colon cancer includes the proximal colon (any primary tumour arising in the cecum, ascending colon, hepatic flexure, or transverse colon), the distal colon (any primary tumour arising in the splenic flexure, descending colon, or sigmoid colon), and cases with an unspecified site. Rectal cancer includes any primary tumour arising in the rectum or rectosigmoid junction” |
|  | e) | Provide details of ethics committee approval and participant informed consent, if relevant | 11 | “All samples for the exposure and outcome datasets were sourced with ethically and obtained approval from the appropriate ethical committee(s)” |
| 5 | **Assumptions** | Explicitly state the three core IV assumptions for the main analysis (relevance, independence and exclusion restriction) as well assumptions for any additional or sensitivity analysis | 6 | “MR uses genetic variants, typically single nucleotide polymorphisms (SNPs), which under specific assumptions can be used in an instrumental variable framework to obtain causal estimates. The three core assumptions are: (i) the genetic variant(s) must be associated with the exposure; (ii) there are no confounders of the association between the genetic variant(s) and the outcome; and (iii) the genetic variant(s) is/are only associated with the outcome via an association with the exposure (14). In addition to these core assumptions, additional assumptions, such as exposure and outcome data being obtained from non-overlapping populations from the same underlying population in summary-level MR, also exist (18); see Sanderson et al. (19) for a detailed overview of MR assumptions.” |
| 6 | **Statistical methods: main analysis** | Describe statistical methods and statistics used |  |  |
|  | a) | Describe how quantitative variables were handled in the analyses (i.e., scale, units, model) | 7 | “Results are given as odds ratio (OR) of CRC risk per standard deviation (SD) higher expression of the gene in CD4+ T cells” |
|  | b) | Describe how genetic variants were handled in the analyses and, if applicable, how their weights were selected |  |  |
|  | c) | Describe the MR estimator (e.g. two-stage least squares, Wald ratio) and related statistics. Detail the included covariates and, in case of two-sample MR, whether the same covariate set was used for adjustment in the two samples | 5 | “(…) MR was then performed to calculate Wald-Ratios.” |
|  | d) | Explain how missing data were addressed | 5-7 | “To account for the lack of allele frequency information in the Fernandez-Rozadilla et al. (2023) summary statistics, we used the most conservative approach to harmonize the data – the harmonize function on the TwoSampleMR (version 0.5.7) package in RStudio (version 2024.4.2.764) using action 3.”  “(…) we matched SNPs between datasets to obtain rsIDs. The datasets used were the outcome GWASs (Fernandez-Rozadilla et al., 2023 & Huyghe et al., 2019), 1000 Genomes European Population, UK Biobank, and GTEx. This matching process was based on chromosome, genomic position, effect allele and other allele information, and palindromic SNPs were excluded. This method resulted in a minor loss of SNPs, which can be seen in more detail in supplementary table 4.”  “To overcome this problem, we used PLINK2.0 to calculate allele frequencies from the 1000 Genomes European samples. This created a new dataset, which we were then able to match with Fernandez-Rozadilla et al. (2023) by rsID, effect allele and other allele, also accounting for palindromic SNPs. This similarly resulted in a loss of SNPs – refer to supplementary table 5 for further details.” |
|  | e) | If applicable, indicate how multiple testing was addressed | 5 | “Benjamini-Hochberg correction (<0.05) was then applied to account for false discovery rates (FDR).” |
| 7 | **Assessment of assumptions** | Describe any methods or prior knowledge used to assess the assumptions or justify their validity | 6 | Methods to assess assumptions are mentioned within the MR analyses subsection of the methodology. |
| 8 | **Sensitivity analyses and additional analyses** | Describe any sensitivity analyses or additional analyses performed (e.g. comparison of effect estimates from different approaches, independent replication, bias analytic techniques, validation of instruments, simulations) | Figure 1 | See figure 1 |
| 9 | **Software and pre-registration** |  |  |  |
|  | a) | Name statistical software and package(s), including version and settings used | 11-12 | “The bulk of the analyses were performed using RStudio (version 2024.4.2.764) (56). MR analyses were performed using TwoSampleMR (version 0.5.7) (57). Data was manipulated using the following packages: arrow (version 16.1.0), biomaRt (version 2.58.0), data.table (version 1.14.10), dplyr (version 1.1.4), GenomicRanges (version 1.54.1), gwascat (version 2.34.0), gwasvcf (version 0.1.2), rtracklayer (version 1.62.0), stringr (version 1.5.1), tidyr (version 1.3.0) (58–68). Plots were created using ggrepel (version 0.9.5) and ggplot2 (version 3.4.4) (69,70). Allele frequencies were calculated using PLINK2.0 (71). Genetic colocalisation analyses were performed using Cmake (version 3.20.0) and PWCoCo (version 1.0) (28). Priors used in the colocalisation analyses were computed using link in reference (29).” |
|  | b) | State whether the study protocol and details were pre-registered (as well as when and where) |  | Details were not pre-registered. |
|  | **RESULTS** |  |  |  |
| 10 | **Descriptive data** |  |  |  |
|  | a) | Report the numbers of individuals at each stage of included studies and reasons for exclusion. Consider use of a flow diagram |  | Not applicable. All individuals were included at each stage of analysis. |
|  | b) | Report summary statistics for phenotypic exposure(s), outcome(s), and other relevant variables (e.g. means, SDs, proportions) | 5 | “Soskic et al. isolated peripheral mononuclear cells (PMBCs) from blood samples obtained from 119 healthy individuals (56% male; 44% female) of European (“British”) ancestries (mean age 47 ± 15.61)” |
|  | c) | If the data sources include meta-analyses of previous studies, provide the assessments of heterogeneity across these studies |  | Not applicable. |
|  | d) | For two-sample MR:  i.  Provide justification of the similarity of the genetic variant-exposure associations between the exposure and outcome samples  ii.  Provide information on the number of individuals who overlap between the exposure and outcome studies | 6 | “We have used two-sample MR in which summary-level data from two different, non-overlapping GWASs were used to generate causal estimates” |
| 11 | **Main results** |  |  |  |
|  | a) | Report the associations between genetic variant and exposure, and between genetic variant and outcome, preferably on an interpretable scale | 7 | “Results are given as odds ratio (OR) of CRC risk per standard deviation (SD) higher expression of the gene in CD4+ T cells” |
|  | b) | Report MR estimates of the relationship between exposure and outcome, and the measures of uncertainty from the MR analysis, on an interpretable scale, such as odds ratio or relative risk per SD difference | Figure 5 | See figure 5 |
|  | c) | If relevant, consider translating estimates of relative risk into absolute risk for a meaningful time period |  | Not applicable |
|  | d) | Consider plots to visualize results (e.g. forest plot, scatterplot of associations between genetic variants and outcome versus between genetic variants and exposure) |  | See figures 3, 4 and 5 |
| 12 | **Assessment of assumptions** |  |  |  |
|  | a) | Report the assessment of the validity of the assumptions | 6 | Described in the methodology section, under the MR analyses subsection. |
|  | b) | Report any additional statistics (e.g., assessments of heterogeneity across genetic variants, such as *I^2^*, Q statistic or E-value) |  | Not applicable |
| 13 | **Sensitivity analyses and additional analyses** |  |  |  |
|  | a) | Report any sensitivity analyses to assess the robustness of the main results to violations of the assumptions | 8 | See results subsections:  Genetic colocalisation analysis  Shared genetic signals across multiple tissues |
|  | b) | Report results from other sensitivity analyses or additional analyses | 8 | See results subsections:  Genetic colocalisation analysis  Shared genetic signals across multiple tissues |
|  | c) | Report any assessment of direction of causal relationship (e.g., bidirectional MR) | 6 | “(…) and Steiger filtering was used to test the directionality of the relationship.” |
|  | d) | When relevant, report and compare with estimates from non-MR analyses |  | Not applicable |
|  | e) | Consider additional plots to visualize results (e.g., leave-one-out analyses) |  | Figure 4 |
|  | **DISCUSSION** |  |  |  |
| 14 | **Key results** | Summarize key results with reference to study objectives | 8-10 | See discussion. Key literature is referenced. |
| 15 | **Limitations** | Discuss limitations of the study, taking into account the validity of the IV assumptions, other sources of potential bias, and imprecision. Discuss both direction and magnitude of any potential bias and any efforts to address them | 10 | See last paragraph of discussion: “However, several limitations to our analysis exist (…)” |
| 16 | **Interpretation** |  |  |  |
|  | a) | Meaning: Give a cautious overall interpretation of results in the context of their limitations and in comparison with other studies | 8-10 | See discussion. Overall interpretation, including limitations and comparisons with other studies, is given for each candidate gene. |
|  | b) | Mechanism: Discuss underlying biological mechanisms that could drive a potential causal relationship between the investigated exposure and the outcome, and whether the gene-environment equivalence assumption is reasonable. Use causal language carefully, clarifying that IV estimates may provide causal effects only under certain assumptions | 8-10 | Possible biological explanation given for each candidate gene. Example:  “TMEM258 is involved in protein synthesis, folding and trafficking (31). Previous research has demonstrated that dysregulation of TMEM258 expression can lead to endoplasmic reticulum (ER) stress, consequently triggering activation of the unfolded protein response (UPR) (31). UPR is known to be beneficial to CD4+ T cells as it supports differentiation, activation, cytokine production and autophagy (32). This may explain the potential mechanism by which increased TMEM258 expression could reduce CRC risk.” |
|  | c) | Clinical relevance: Discuss whether the results have clinical or public policy relevance, and to what extent they inform effect sizes of possible interventions | 12 | “We were able to provide valuable insights into possible candidate genes for CRC prevention” |
| 17 | **Generalizability** | Discuss the generalizability of the study results (a) to other populations, (b) across other exposure periods/timings, and (c) across other levels of exposure | 10 | “First, we used data from European ancestries which, though we assume are homogenous and therefore satisfy our genetic instrument assumptions, means these results may not be generalizable to other populations.” |
|  | **OTHER INFORMATION** |  |  |  |
| 18 | **Funding** | Describe sources of funding and the role of funders in the present study and, if applicable, sources of funding for the databases and original study or studies on which the present study is based | 11 | Fully described in declarations section. |
| 19 | **Data and data sharing** | Provide the data used to perform all analyses or report where and how the data can be accessed, and reference these sources in the article. Provide the statistical code needed to reproduce the results in the article, or report whether the code is publicly accessible and if so, where | 12 | All code and data used is described in the Availability of data and materials section. |
| 20 | **Conflicts of Interest** | All authors should declare all potential conflicts of interest | 12 | Fully described in declarations section. |

This checklist is copyrighted by the Equator Network under the Creative Commons Attribution 3.0 Unported (CC BY 3.0) license.

1.	Skrivankova VW, Richmond RC, Woolf BAR, Yarmolinsky J, Davies NM, Swanson SA, et al. Strengthening the Reporting of Observational Studies in Epidemiology using Mendelian Randomization (STROBE-MR) Statement. JAMA. 2021;under review.

2.	Skrivankova VW, Richmond RC, Woolf BAR, Davies NM, Swanson SA, VanderWeele TJ, et al. Strengthening the Reporting of Observational Studies in Epidemiology using Mendelian Randomisation (STROBE-MR): Explanation and Elaboration. BMJ. 2021;375:n2233.
