## Supplementary figures and images for "Transcriptome-wide Mendelian randomisation exploring dynamic CD4+ T cell gene expression in colorectal cancer development"

### Supplementary figure 1

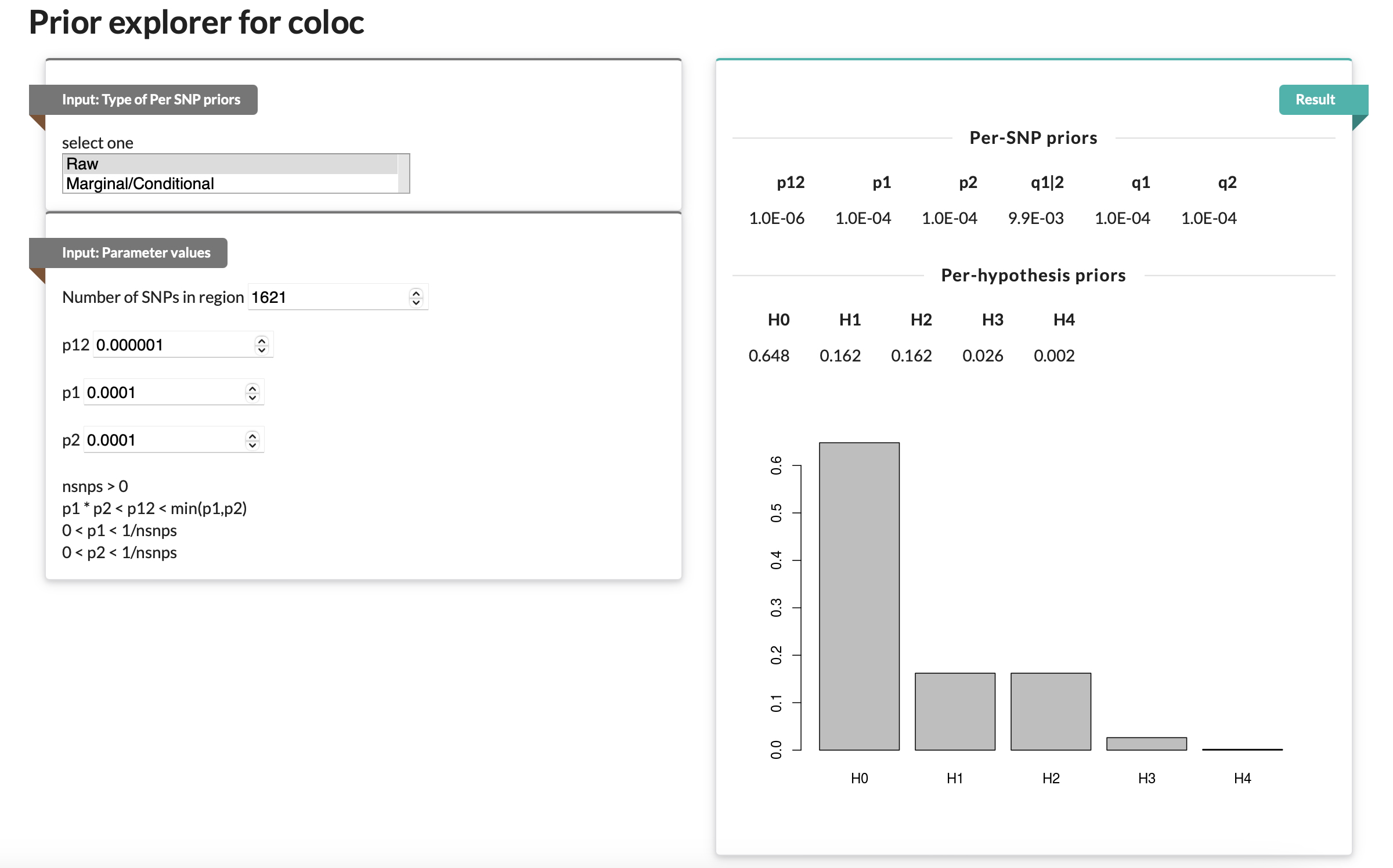
